# Retrospective Analysis of Equity-Based Optimization for COVID-19 Vaccine Allocation

**DOI:** 10.1101/2023.05.08.23289679

**Authors:** Erin Stafford, Dobromir Dimitrov, Rachel Ceballos, Georgina Campelia, Laura Matrajt

## Abstract

Marginalized racial and ethnic groups in the United States were disproportionally affected by the COVID-19 pandemic. To study these disparities, we construct an age-and-race-stratified mathematical model of SARS-CoV-2 transmission fitted to age-and-race-stratified data from 2020 in Oregon and analyze counter-factual vaccination strategies in early 2021. We consider two racial groups: non-Hispanic White persons and persons belonging to BIPOC groups (including non-Hispanic Black persons, non-Hispanic Asian persons, non-Hispanic American Indian or Alaska Native persons, and Hispanic or Latino persons). We allocate a limited amount of vaccine to minimize overall disease burden (deaths or years of life lost), inequity in disease outcomes between racial groups (measured with five different metrics), or both. We find that, when allocating small amounts of vaccine (10% coverage), there is a trade-off between minimizing disease burden and minimizing inequity. Older age groups, who are at a greater risk of severe disease and death, are prioritized when minimizing measures of disease burden, and younger BIPOC groups, who face the most inequities, are prioritized when minimizing measures of inequity. The allocation strategies that minimize combinations of measures can produce middle-ground solutions that similarly improve both disease burden and inequity, but the trade-off can only be mitigated by increasing the vaccine supply. With enough resources to vaccinate 20% of the population the trade-off lessens, and with 30% coverage, we can optimize both equity and mortality. Our goal is to provide a race-conscious framework to quantify and minimize inequity that can be used for future pandemics and other public health interventions.

## 1 Introduction

The COVID-19 pandemic has highlighted and exacerbated the inequities of the health care system in the United States (US) and other countries in the world. In the US, over the course of the pandemic, there have been significant inequities in cases, hospitalizations, and deaths according to race and ethnicity, with those in marginalized communities carrying the most burden of the pandemic. In the state of Oregon, racially marginalized communities in every age-group have experienced disproportionate mortality rates. For example, the Non-Hispanic (NH) American Indian or Alaska Native (AIAN) population had the highest risks of COVID-19 infection, hospitalization, and death and were 2.7, 3.6, 3.2 times more likely to be infected, hospitalized, or die when compared to a white person of the same age in 2020 [1]. Moreover, the NH black or African-American persons, NH Asian persons, NH AIAN persons, and Hispanic or Latino persons aged 20-59 have experienced mortality rates 5.7 times higher than their white counter parts, those aged 60-69 have experienced 4 times higher rates, and individuals aged 0-19 and 70+ have experienced mortality rates that are 1.8 times higher. Data has shown similar disparities nationally [2]. Studies have shown that this is due, in part, to systemic racism and inequities leading to differences in comorbidities, access to health care, and occupation [2]–[21]. For example, BIPOC persons are more likely to be employed in front-line work, leading to more exposure, and decreasing the effectiveness of mitigation strategies like shelter-in-place for these groups [22]–[26].

In the summer of 2020, the National Academies of Science, Medicine and Engineering (NASEM) and the World Health Organization (WHO) released frameworks for equitable vaccine allocation [27], [28]. However, allocating the vaccine in an equitable way proved to be difficult, as there were many obstacles, including differences in vaccine perception [29] and inequities in access [22], [30], resulting in large differences in vaccination rates between racial groups, particularly during the first few months of the vaccination campaigns [31]–[34]. These inequities have further compounded disparities in COVID-19 outcomes [35]. Hence, the need to create health policies, including vaccination strategies, that promote equity through the use of targeted, community-based interventions is increasingly evident [5], [22], [36], [37].

Previous work addressing vaccine prioritization for COVID-19 vaccines has predominantly been age-centered (e.g. [38]–[44]) with only a few studies including social variables [45]. Most of these studies evaluate vaccination success as the reduction of the overall disease burden (mortality, hospitalizations, or infections) without accounting for projected equity gaps. A few studies included inequity outcomes mostly focusing on geographic distribution of vaccines among countries, states, or affected regions [46], [47], and only a handful discuss reducing inequities in access or disparities in outcomes between racial groups [48].

In the present work, we explore a race conscious, as opposed to colorblind, approach to determine mathematically optimal vaccine allocation strategies that not only minimize overall disease burden (mortality or years of life lost, YLLs) but also minimize the inequity in COVID-19 outcomes between racial groups, which we measure using several metrics. To that end, we develop an age-and-race-stratified mathematical model, and apply optimization algorithms, which account for both, disease burden and equity, to determine the optimal distribution of available vaccine doses across age and race groups under different disease and equity metrics. We retrospectively analyze data from Oregon for the first four months of 2021, when vaccine supplies were extremely limited and SARS-CoV-2 was relatively stable and explore counterfactual vaccine allocations. To be clear, we are not suggesting allocating vaccine based on race, but given the fact that the data itself is race-stratified, we use race as an incomplete proxy for systemic inequities, such as disparities in social determinants of health (SDOH), affecting BIPOC communities. Nevertheless, we recognize that considering race alone does not grant us full understanding of the systemic inequities caused by multiple axes of oppression [49] and that no mathematical model or vaccine allocation strategy alone could be sufficient to resolve the profound and complex problem of racism and centuries-long systemic inequities experienced in the US. Policy makers are faced with extreme pressure during a public health emergency, such as a pandemic, where they need to make decisions affecting all aspects of our lives. Our hope is to provide a framework to learn from the inequities observed during the COVID-19 pandemic, and that other public health interventions, present and future, might benefit from a quantitative approach addressing both disease burden and inequity reduction. Our analysis suggests that when vaccine supply is very limited, there is a trade-off between diminishing overall mortality and reducing inequity resulting in significantly high, and likely unacceptable, levels of burden in one or the other. However, this trade-off lessens as more vaccine becomes available.

### 1.1 The mathematical model

We construct a deterministic mathematical model of SARS-CoV-2 transmission and vaccination. The structure of our model accommodates the available case data from the state of Oregon. This data includes information about hospitalization and death after infection as well as demographic information about age and race or ethnicity. Although much of the inequity in COVID-19 outcomes between racial or ethnic groups is due to systemic disparities in SDOH such as access to healthcare, income, and education, this information is not regularly collected along with case data. Therefore, we use the race and ethnicity variable as a proxy for SDOH and these systematic disparities to gain insight into counteracting or preventing inequities in resource allocation and disease outcomes [50].

In our model, individuals can be either unvaccinated or vaccinated. Once infected, susceptible individuals become latent, meaning they are infected but not yet infectious. Once the latency period has ended, individuals may become either symptomatic or asymptomatic. All asymptomatic individuals eventually recover, but symptomatic individuals may become hospitalized before recovering or dying (Figure 1A).

**Figure 1:**
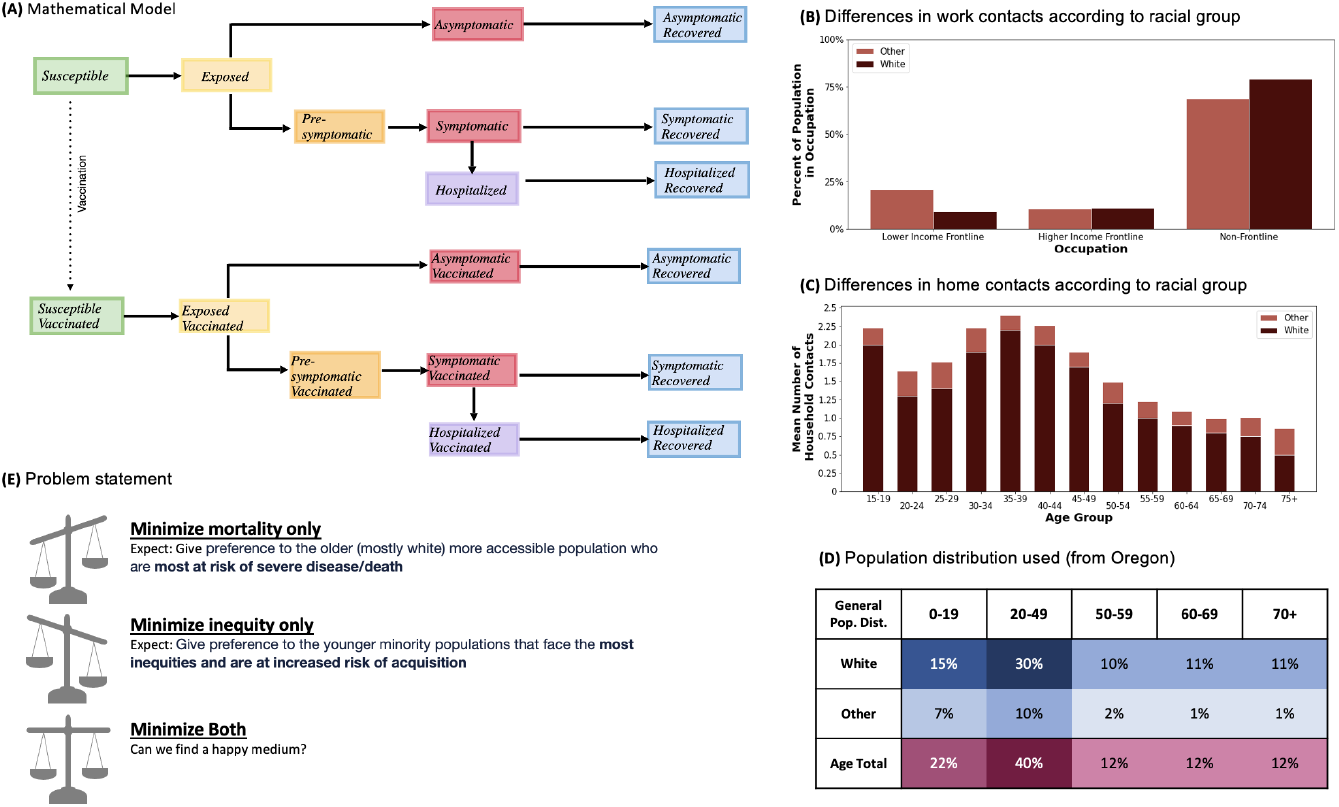
Model description and demographic information. This figure includes the model diagram (A), the differences in occupation type according to racial group [56] (B), the differences in home contacts according to racial group [54] (C), the population distribution of the population under study (D), and the problem statement (E).

We stratified the population into five age groups: children (aged 0-19), adults aged 20-49, adults aged 50-59, adults aged 60-69 years old, and adults aged 70 and older. We chose not to vaccinate children in our model as vaccines were not available for children at the beginning of 2021. We consider two racial groups: non-Hispanic white persons (referred to as white) and persons belonging to other racial or ethnic groups as preestablished in the fitted data (including non-Hispanic black or African-American persons, non-Hispanic Asian persons, non-Hispanic American-Indian or Alaska-Native persons, and Hispanic or Latino persons), referred to as BIPOC below. For each age group, the BIPOC racial group was computed as a weighted average of the racial/ethnic groups mentioned above, reflecting the composition of the population in Oregon in 2020-2021 according to census data for the white and BIPOC groups [51] (Figure 1D).

Mortality and hospitalization rates by age were estimated from published sources [52] and [53]. Using data from Oregon, we computed per-capita mortality rate ratios between the BIPOC and white groups for each age group and use these ratios to estimate the differences in health outcomes between racial groups.

In our model, individuals come into contact with each other in four locations: home, school, work, and community. Using data given in [54], [55] and [56], we adapted the contact matrices from [57] for home and work, respectively, to reflect different contact patterns across racial groups. Figure 1 shows the resulting contact patterns stratified by race used to adapt the work (B) and home (C) contacts. We assume community and school contacts are not different for racial groups, but these contacts are proportional to the population size of each group. The overall contact matrix is found by summing the location-specific contact matrices.

Following the ideas of [58], [59], we consider three vaccine effects: reduction on the susceptibility to infection (set to 70%), reduction on symptomatic infection (set to 66%) and reduction on hospitalization and death (set to 90%). These values reflect the vaccine effectiveness estimated as of January 2021 [58], [60]–[63]. We assume that natural immunity, which is completely effective, and vaccine-induced immunity last for the entirety of the simulated period of four months with no new strains of COVID-19 introduced during that time.

### 1.2 The optimization problem

Our goal is to distribute available vaccines to the four, adult age-groups in the two racial groups aiming to minimize different objective functions including measures of disease burden (mortality and YLLs), measures of inequity (Table 1), and joint disease burden and inequity measures. For the combined measures, the inequity measure is weighted to match the scale of the measure of disease burden. To calculate YLLs, the number of deaths in each group is multiplied by the average remaining life expectancy in that group. Therefore, YLLs assign higher values to the lives lost in the younger populations [64], [65].

**Table 1:**
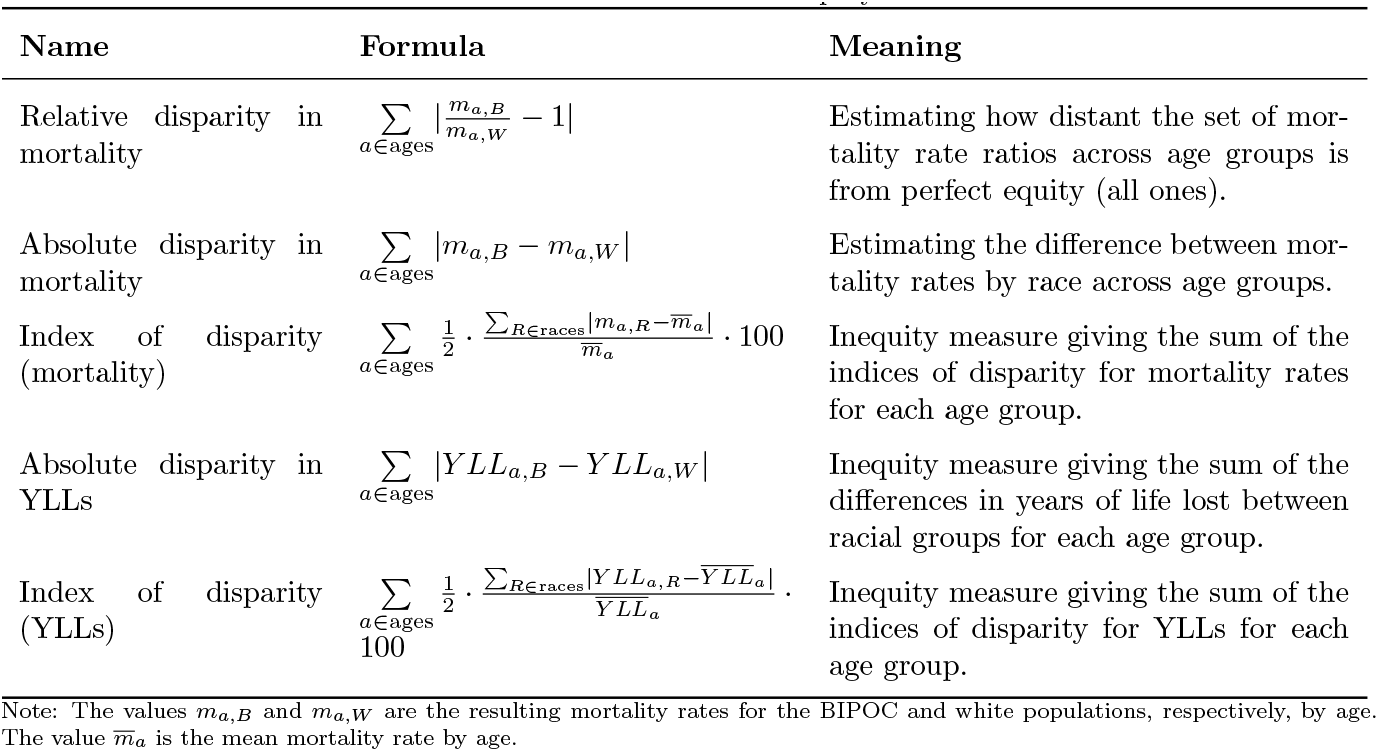
Measures of Inequity

We consider five measures of inequity, described in Table 1. These include a relative and absolute disparity in mortality rates between racial groups calculated on age-group basis as well as the absolute disparity in YLLs between racial groups. We also include indices of disparity in mortality and YLLs based on race/ethnicity [66]. These indices are modified coefficients of variation and are commonly used to study disparities in disease outcomes [67], [68].

For our optimization routine, we develop a coarse global search algorithm to find the vaccination strategy that minimizes a given objective function with a fixed amount of resources (vaccine doses) over a time frame of four months. Vaccination strategies are represented by vectors of the proportion of each group to be vaccinated (ranging from 0 to 1). Similar to [38], we performed the optimization routine in two steps. First, we generate 10,000 random vaccination vectors and evaluate the objective function using each. We select the 20 best vectors and use them, together with four educated-guesses, as starting points for three constrained optimization algorithms: trust-region constrained, SLSQP, and adapted Nelder-Mead. We report the optimal allocation strategy to be the best solution among the 24 solutions calculated

We focus on a scenario where vaccine availability is very constrained similar to the situation in the United States in January 2021. We explored scenarios with enough vaccine to cover 10%, 20%, or 30% of the entire population.

## 2 Results

### 2.1 Model fitting

We calibrate our model to the number of deaths reported in Oregon in 2020 by fitting the percentage of contacts maintained when social distancing policies were implemented as well as the rates of asymptomatic infection by age. Given our interest in the disparities in severe COVID-19 outcomes between different races, we also fit to the mortality rates ratios between the white and BIPOC populations in each age group (Figure 2, Table 2).

**Table 2:** Comparison of deaths from model fitting to data from Oregon.

| Deaths Per-Group | 0-19 | 20-49 | 50-59 | 60-69 | 70+ |
| --- | --- | --- | --- | --- | --- |
| Data: White | 0 | 17 | 49 | 173 | 1130 |
| Data: Other | 1 | 33 | 55 | 76 | 170 |
| **Data Total: 1704** |  |  |  |  |  |
| Model: White | 1 | 11 | 50 | 253 | 968 |
| Model: Other | 2 | 24 | 62 | 132 | 202 |
| **Model Total: 1704** |  |  |  |  |  |

**Figure 2:**
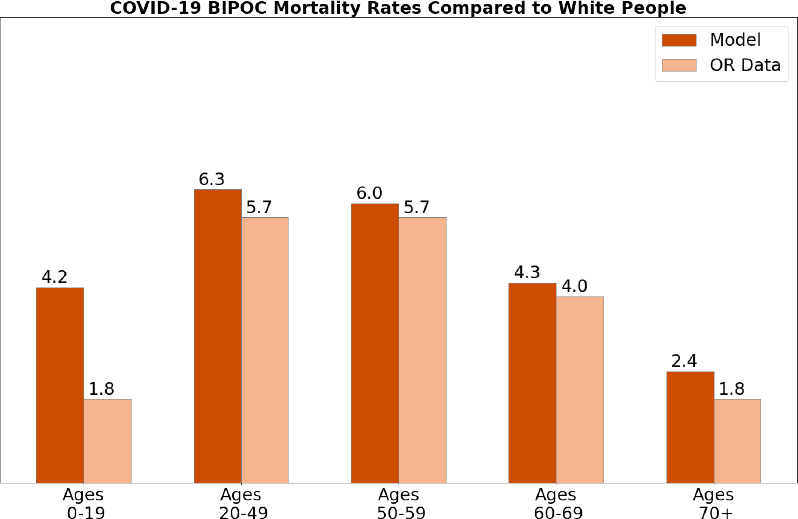
Mortality rate ratios from model fitting compared to data.

### 2.2 Allocating vaccines

We start with a scenario in which vaccine supply is very limited by assuming that available vaccine doses in the beginning of 2021 are enough to cover 10% of the population. We simulate a baseline case where vaccines are randomly allocated to serve as a basis for comparison to the allocation strategies found through optimization, referred as optimal strategies below (Figure 3). In the baseline case, the burden of disease, measured by the number of deaths or YLLs, is concentrated in the oldest age-groups. Moreover, individuals aged 70+ from the BIPOC group die at 2.4 times higher rates than their white counterparts (Figure 3C). The most extreme racial inequities in burden of disease, however, are in the young and middle-aged adult groups, where there is a maximum relative difference in mortality of 6.2 in those aged 20-49 years old and the highest index of disparity (134) in those aged 50-59 (Figure 3D).

**Figure 3:**
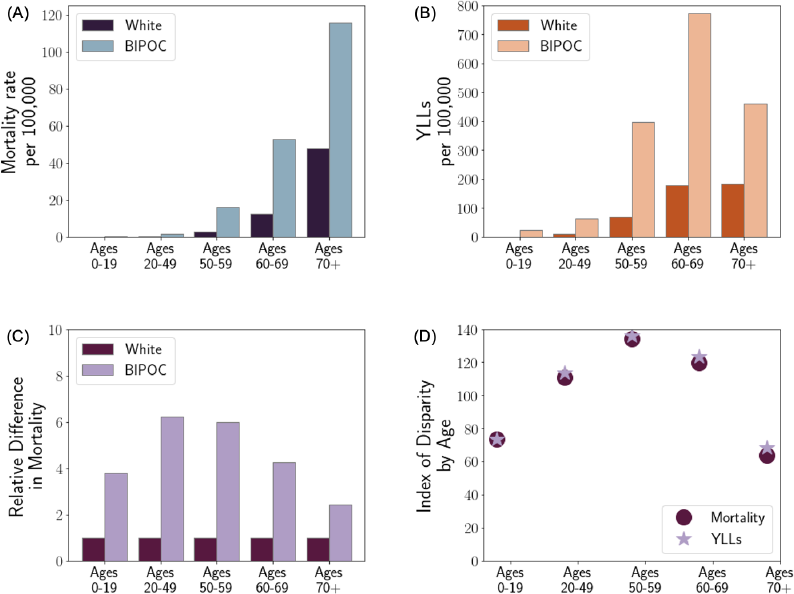
Outcomes of the baseline allocation strategy.

We then allocate vaccines by minimizing each of the objective functions defined above. When minimizing only measures of disease burden (deaths or YLLs) or only measures of inequity, the optimal allocation strategy is to prioritize either the older population (who are most at risk of severe disease and death) or the younger, BIPOC population (who face the most inequities), respectively (Figure 4). To minimize combinations of measures, the optimal strategy is to strike a balance between the objectives. For example, to minimize both deaths and relative inequity in mortality, a significant amount of the available vaccine (about 73%) is allocated to the older population to reduce mortality, and the remaining vaccine (about 27%) is allocated to the younger BIPOC population to reduce inequity. In this case, neither population is completely prioritized over the other.

**Figure 4:**
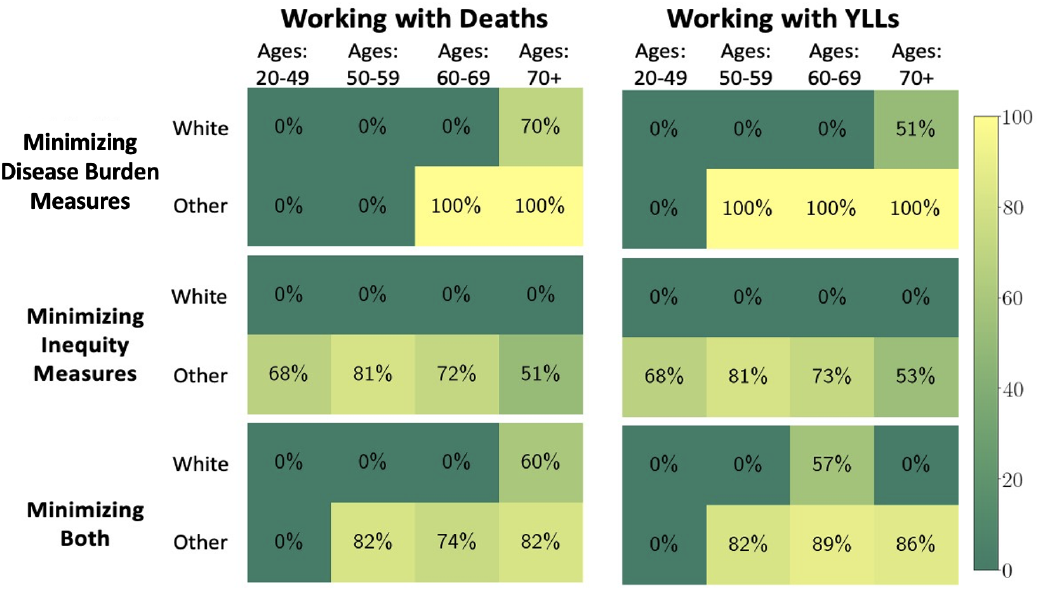
Vaccine allocation with resources to vaccinate 10% of the population. The first column in this figure contains the results of minimizing deaths (row 1), relative disparity in deaths (row 2), and deaths and relative disparity in mortality (row 3). The second column in this figure contains the results of minimizing YLLs (row 1), absolute disparity in YLLs (row 2), and YLLs and the absolute disparity in YLLs.

All of our optimal vaccine-allocation strategies improve COVID-19 outcomes, in both disease burden and inequity measures, when compared to the baseline scenario (Figure 5). There is, however, a trade-off. Minimizing only measures of disease burden results in vaccine allocation strategies with larger inequity and vice versa. This holds for any optimization based on a single metric (disease burden or inequity) and for all the inequity metrics considered (Table 1, Figure 5A vs Figure 5B). For example, minimizing overall deaths reduces mortality by 61% from the baseline scenario, but minimizing relative inequity in mortality reduces overall mortality by only 16%.

**Figure 5:**
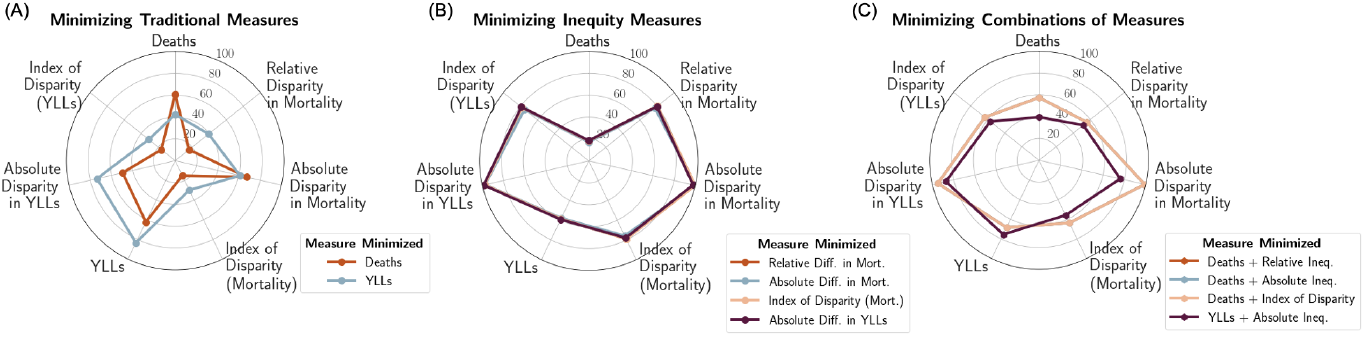
Summary of COVID-19 outcomes when allocating vaccine to 10% of the population. The vaccine allocation strategies that minimize measures of disease burden (A), inequity measures (B), and combinations of measures (C) are evaluated. The results are reported as the percent averted from the base (random allocation) case.

The optimal allocation strategies that minimize different metrics of inequity yield very similar outcomes (Figure 5B). When comparing between measures of disease burden, minimizing only YLLs is more equitable than minimizing only deaths and results in a greater percentage of inequity averted for four of the five measures of inequity (Figure 5A). As expected, minimizing YLLs leads to more deaths (42% deaths averted when compared to baseline versus 61% when minimizing deaths), but significantly decreases the number of YLLs (83% YLLs averted versus 62% when minimizing deaths).

Allocation strategies minimizing combined measures of disease burden and inequity produce middle-ground solutions improving both. For instance, when the overall number of deaths is minimized along with any of the measures of inequity in mortality, mortality is reduced by 57% and the relative disparity in mortality is reduced by 56% from the baseline. This results in 4 percentage points less deaths averted than when minimizing mortality alone and 25 percentage points less inequity averted than when minimizing inequity alone (Figure 5C). Minimizing both YLLs and measures of inequity based on YLLs results in a 75% reduction in YLLs, and an 87% reduction in absolute disparity in YLLs when compared to baseline. This shows a similar trade-off when compared to single-measure optimizations minimizing YLLs (83% reduction in YLLs) or inequity in YLLs only (99% reduction in absolute disparity in YLLs).

Our analysis highlights the importance of accounting for both disease burden and inequity, when vaccine supply is limited. As more vaccine doses become available, the trade-off between equity and mortality lessens. With enough resources to vaccinate 20% of the population, there are more vaccination strategies significantly reducing both equity and mortality simultaneously, and combined optimization yields similar impact in minimizing both outcomes as minimizing each one individually. For example, the optimal allocation strategy minimizing both overall deaths and relative inequity in mortality reduces mortality by 83% and inequity by 71% from the baseline scenario. This is comparable to the 89% and 85% reduction in deaths and inequity in mortality obtained when minimizing each alone (Figure 6). With a further increase of available resources (enough to vaccinate 30% of the population) the difference between the combined and single-measure optimizations is almost entirely eliminated. In this case, minimizing both overall deaths and relative inequity in mortality results in comparable reduction in mortality with minimizing only deaths (94% vs 97%) while preserving the majority of the inequity reduction (78%) compared to scenarios minimizing only inequity (85%).

**Figure 6:**
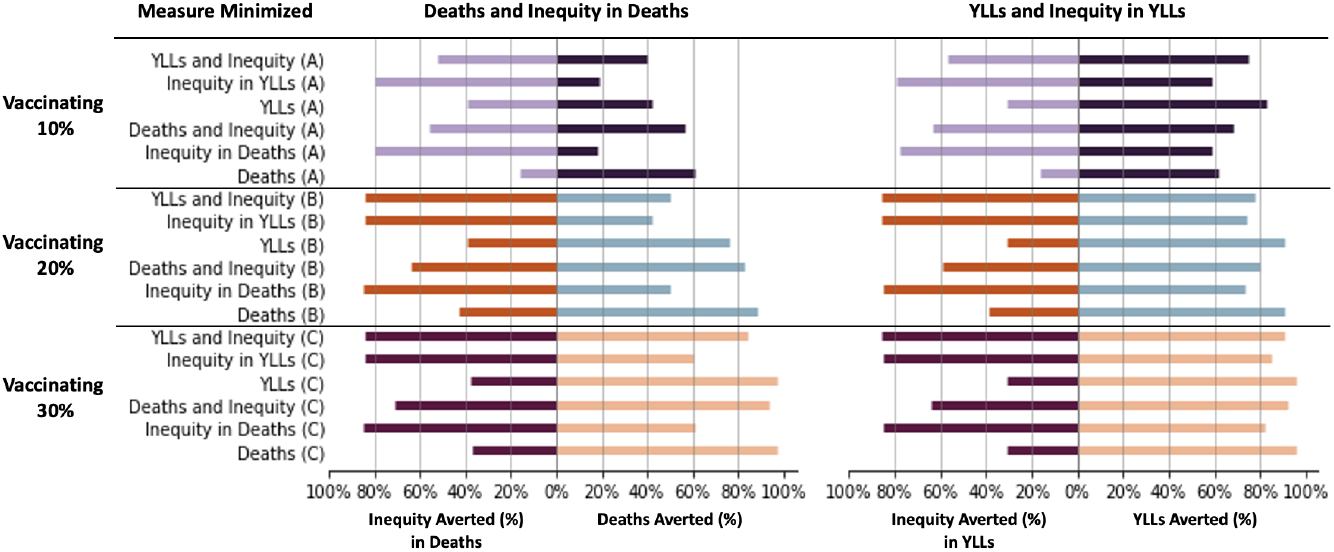
Performance of optimal vaccination strategies for six minimization objectives when there is enough vaccine to cover 10%, 20% or 30% of the population. Each strategy is evaluated for four metrics: overall deaths, YLLs, inequity in deaths (measured by the index of disparity in mortality), and inequity in YLLs (measured by the index of disparity in YLLs). Performance is reported as the percent averted compared to the baseline scenario with random vaccination.

## 3 Discussion

During late 2020 and early 2021, different voices proposed vastly different vaccine allocation strategies [69]. Some argued that protecting the older population, which is mostly white and at the highest risk for severe disease, needed to be prioritized [70], [71]. Others argued that vaccines needed to be allocated to communities experiencing the greatest inequities in access and disparities in outcomes, usually younger BIPOC populations, working more often in essential occupations [72]–[75]. Here, we are not advocating for the allocation of vaccines based on race or ethnicity. We instead use race and ethnicity as a proxy for social determinants of health such as access to healthcare and occupation for which data is not usually collected along with case information. While racial identity is a poor proxy for analyzing and addressing systemic inequities in vaccine allocation and disparities in YLLs or mortality, it allows a race conscious data-driven analysis that can be incorporated into broader societal decisions about how to best allocate vaccine in urgent conditions of scarcity, such as those experienced early in the SARS-CoV-2 pandemic. For example, the results when minimizing inequity support prioritizing marginalized communities. This could be interpreted as implementing mitigation strategies such as setting up more vaccination clinics in underserved communities, using community-engaged public health approaches, or first vaccinating frontline workers. It also highlights the urgent need to collect better data regarding social variables, so that further analyses can be refined.

In the present work, we used an age-and-race-structured mathematical model to study counterfactual scenarios of COVID-19 vaccine allocation in early 2021 in the United States, when very limited amounts of vaccine were available. Our results are not meant to be construed as a critique of the COVID-19 vaccine allocation during the pandemic, but rather as a theoretical exercise to quantify equity in public health resource distribution. Indeed, our results are aligned with the prioritization followed by most regions worldwide when vaccine availability was extremely low, where older adults were given vaccine first to minimize mortality. To better understand the effect of different vaccination policies on inequity, we analyzed five measures of inequity. Our results suggest that when vaccine is very limited, there is a trade-off between minimizing inequity and minimizing overall mortality (or YLLs). If minimizing overall mortality, vaccine was allocated to the oldest population, with those in racially marginalized older groups being prioritized. In contrast, when minimizing inequity, irrespective of the metric used, vaccine was allocated to those in the, usually younger, BIPOC group. This resulted in additional deaths in older age groups, including in BIPOC communities. This trade-off, however, lessens as more resources become available, and our optimization work shows that it is possible to minimize both mortality and inequity if vaccine is allocated optimally.

The trade-off between equity and mortality is exaggerated due to the nature of COVID-19 disease profile: the burden of severe disease and mortality is concentrated in the oldest population, which is disproportionately white due to racism leading to inequities in access to care and disparities in morbidity/mortality at baseline. As vaccines become more readily available, the potential to address both age-and-racial/ethnic inequities mathematically, becomes more balanced. Translation of this potential then needs to shift to evaluation of systemic biases that impact vaccination allocation policies, implementation strategies, and conscious and unconscious bias that impact vaccine distribution. In that sense, our work suggests that for other infectious diseases which do not have a strong age-dependent mortality pattern (e.g., mpox or HIV), resources can be allocated to reduce overall disease burden and inequity simultaneously, even when they are scarce.

Our work, like any mathematical model, is subject to limitations. First, due to data limitations, we use race and ethnicity as a proxy for factors that lead to differences in health outcomes such as social determinants of health and racism in society. Therefore, our results may serve as an indication of which groups to prioritize for vaccine distribution, but do not tell us which factors have the most influence. For example, we are not able to determine if individuals with low access to healthcare should be prioritized over those in essential occupations. Furthermore, we only included two racial/ethnic groups. However, the BIPOC group was calculated to match the proportions in Oregon of each of the five racial/ethnic groups defined in [2], and we believe that the inclusion of more racial/ethnic groups will not significantly alter the results. While these proportions closely match the national average, the optimal vaccination strategies could potentially be different in other cities where the composition of the population is very different.

In the first few months of 2021, the majority of the population was unvaccinated, SARS-CoV-2 reinfections were very uncommon, and the virus was relatively stable. Therefore, we did not include waning immunity from previous infections, or from vaccinations, or booster vaccinations. Furthermore, we currently vaccinate all individuals simultaneously at the beginning of our simulation. This allows us to compare different vaccination strategies without additional confounders (e.g., vaccination rates). Implementing a vaccination campaign would be more realistic but would require us to also optimize the order in which individuals are vaccinated. We quantified inequity in five different ways and combined them with mortality or YLLs, but other measures or combinations might be more appropriate. In particular, using Pareto optimization instead of adding these measures might be a better way to explore the trade-offs between reduction in overall mortality and minimizing inequitable outcomes. Much of the inequity in disease outcomes is the result of profound systemic inequities that cannot be overcome through vaccine allocation alone. Therefore, absolute equity is not achievable.

Our results show that public health decisions in the midst of a pandemic are intrinsically difficult and that trade-offs might be unavoidable when public health crises arise in a society that is inequitable and rooted in racially biased systems at baseline. Indeed, decision-makers need to consider societal, economic, and political factors to understand and address the trade-offs that arise. While counteracting racial inequities and underlying racism is complex and can be emotionally charged, our work may help to provide a quantitative framework to measure the impact of public health interventions on equity. Specifically, it offers quantitative methodology to evaluate strategies for counteracting racial/ethnic inequities in the distribution of resources and in outcomes. We hope that this framework will help inform future discussions about how to equitably protect those at highest risk of harm in public health emergencies.

## Data Availability

We acknowledge the data-sharing agreement with the Oregon Health Authority (DUA220114) that allowed us access the COVID-19 data from Oregon.

## Acknowledgements

This work was partially supported by grants from the National Institutes of Health (UM1AI068635 and UM1AI068617). L.M. and D.D. were also supported by a grant from Centers for Disease Control and Prevention (NU38OT000297-02) through their cooperative agreement with the Council of State and Territorial Epidemiologists. This material is based upon work supported by the National Science Foundation under Grant No.2210382. We acknowledge the data-sharing agreement with Oregon Health Authority (DUA220114) that allowed us access the COVID-19 data from Oregon, and we would like to thank Erik Everson for his help with accessing this data. The content is solely the responsibility of the authors and does not necessarily represent the official views of any of the funders of this work.

