## Supplementary Information for "Retrospective Analysis of Equity-Based Optimization for COVID-19 Vaccine Allocation"

[a]Department of Applied Mathematics, University of Washington, Seattle, WA
[b]Vaccine and Infectious Disease Division, Fred Hutchinson Cancer Center, Seattle, WA
[c]Cancer Control and Population Sciences, Huntsman Cancer Institute, Salt Lake City, UT
[d]Department of Family and Preventive Medicine, University of Utah, Salt Lake City, UT
[d]Department of Bioethics and Humanities, University of Washington, Seattle, WA
[*]Corresponding Author

### Supplementary Information

The model equations for the deterministic age-structured mathematical model of SARS-CoV-2 transmission stratified by both age and race for the unvaccinated population are

$$\dot{S}_i = -\lambda S_i, \tag{1}$$
$$\dot{E}_i = \lambda S_i - \gamma_E E_i, \tag{2}$$
$$\dot{A}_i = \gamma_E(1-a_i)E_i - \gamma_A A_i, \tag{3}$$
$$\dot{P}_i = \gamma_E a_i E_i - \gamma_P P_i, \tag{4}$$
$$\dot{I}_i = \gamma_P P_i - (1-h_i)\gamma_I I_i - \sigma h_i I_i, \tag{5}$$
$$\dot{H}_i = \sigma h_i I_i - \gamma_H H_i, \tag{6}$$
$$\dot{R}_i = (1-h_i)\gamma_I I_i, \tag{7}$$
$$\dot{RA}_i = \gamma_A A_i, \tag{8}$$
$$\dot{RH}_i = (1-d_i)\gamma_H H_i. \tag{9}$$
$$\tag{10}$$

For the vaccinated population, the model equations are

$$\dot{Sv}_i = -\theta_V \lambda Sv_i, \tag{11}$$
$$\dot{Ev}_i = \theta_V \lambda Sv_i - \gamma_E Ev_i, \tag{12}$$
$$\dot{Av}_i = \gamma_E(1-\phi_V a_i)Ev_i - \gamma_A Av_i, \tag{13}$$
$$\dot{Pv}_i = \gamma_E \phi_V a_i Ev_i - \gamma_P Pv_i, \tag{14}$$
$$\dot{Iv}_i = \gamma_P Pv_i - (1-\rho_V h_i)\gamma_I Iv_i - \sigma\rho_V h_i Iv_i, \tag{15}$$
$$\dot{Hv}_i = \sigma\rho_V h_i Iv_i - \gamma_H Hv_i, \tag{16}$$
$$\dot{Rv}_i = (1-h_i)\gamma_I Iv_i, \tag{17}$$
$$\dot{RAv}_i = \gamma_A Av_i, \tag{18}$$
$$\dot{RHv}_i = (1-\delta_V d_i)\gamma_H Hv_i. \tag{19}$$
$$\tag{20}$$

The force of infection is

$$\lambda = \sum_{i=1}^{10} \frac{r_i C\beta}{N_i}\left[r_A(A_i + \psi_V Av_i) + r_P(P_i + \psi_V Pv_i) + (I_i + \psi_V Iv_i) + r_H(H_i + \psi_V Hv_i)\right]. \tag{21}$$

The age-and-race-stratified contact matrix, $C$, is adapted from the age-specific contact matrix for the US given in [1], and we calculate $\beta$ by assuming $R_0 = 3$. The parameters used for this model are described in Table S1.

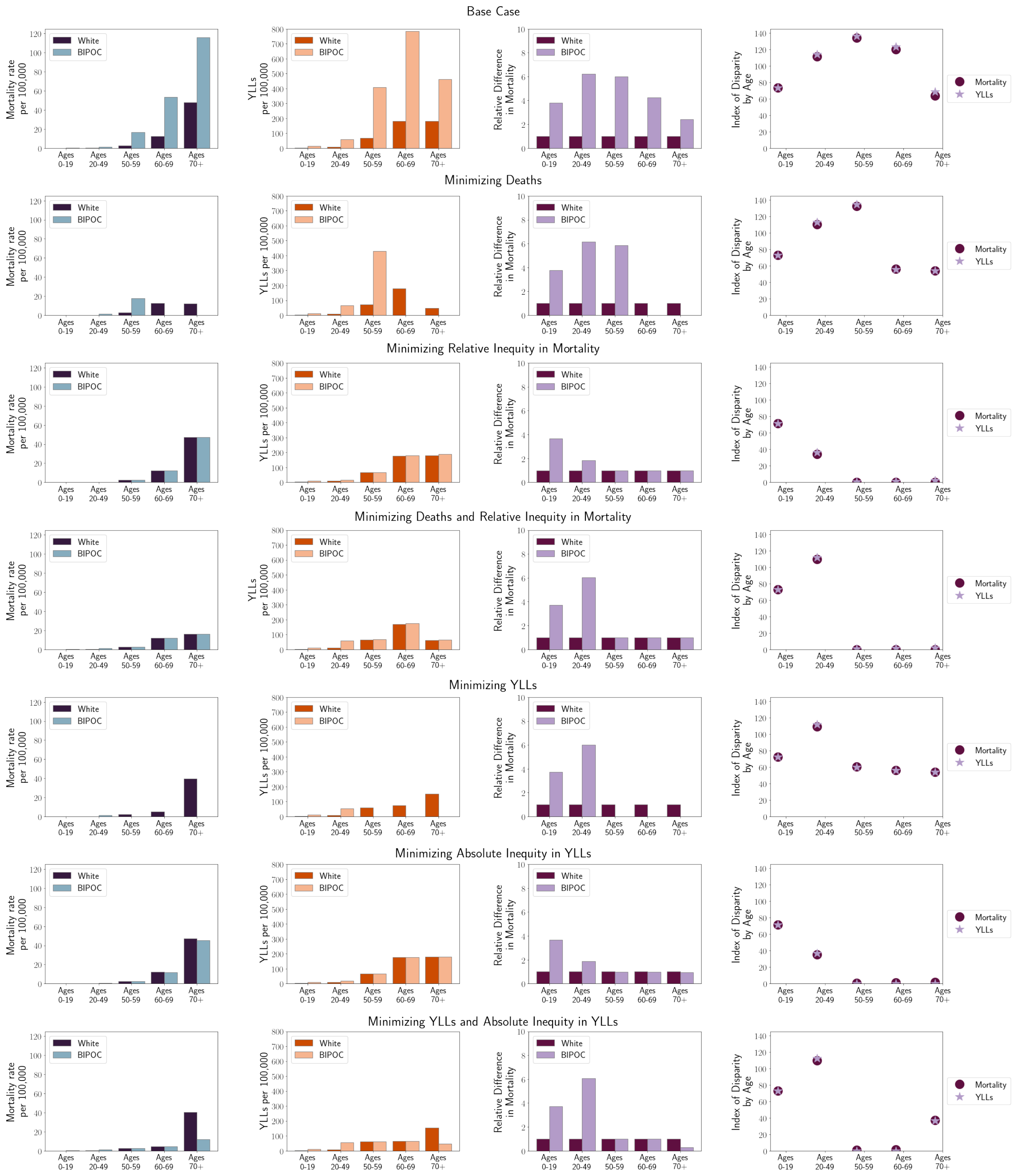


Figure 1: Summary of COVID-19 outcomes by-age when allocating vaccine to 10% of the population. The vaccine allocation strategies that minimize measures of disease burden (rows 2 and 5), inequity measures (rows 3 and 6), and combinations of measures (rows 4 and 7) are evaluated for comparison to the base case (row 1). Outcomes including mortality per 100,000 (column 1), YLLs (column 2), relative disparity in mortality (column 3), and the index of disparity in mortality and YLLs (column 4) are shown.

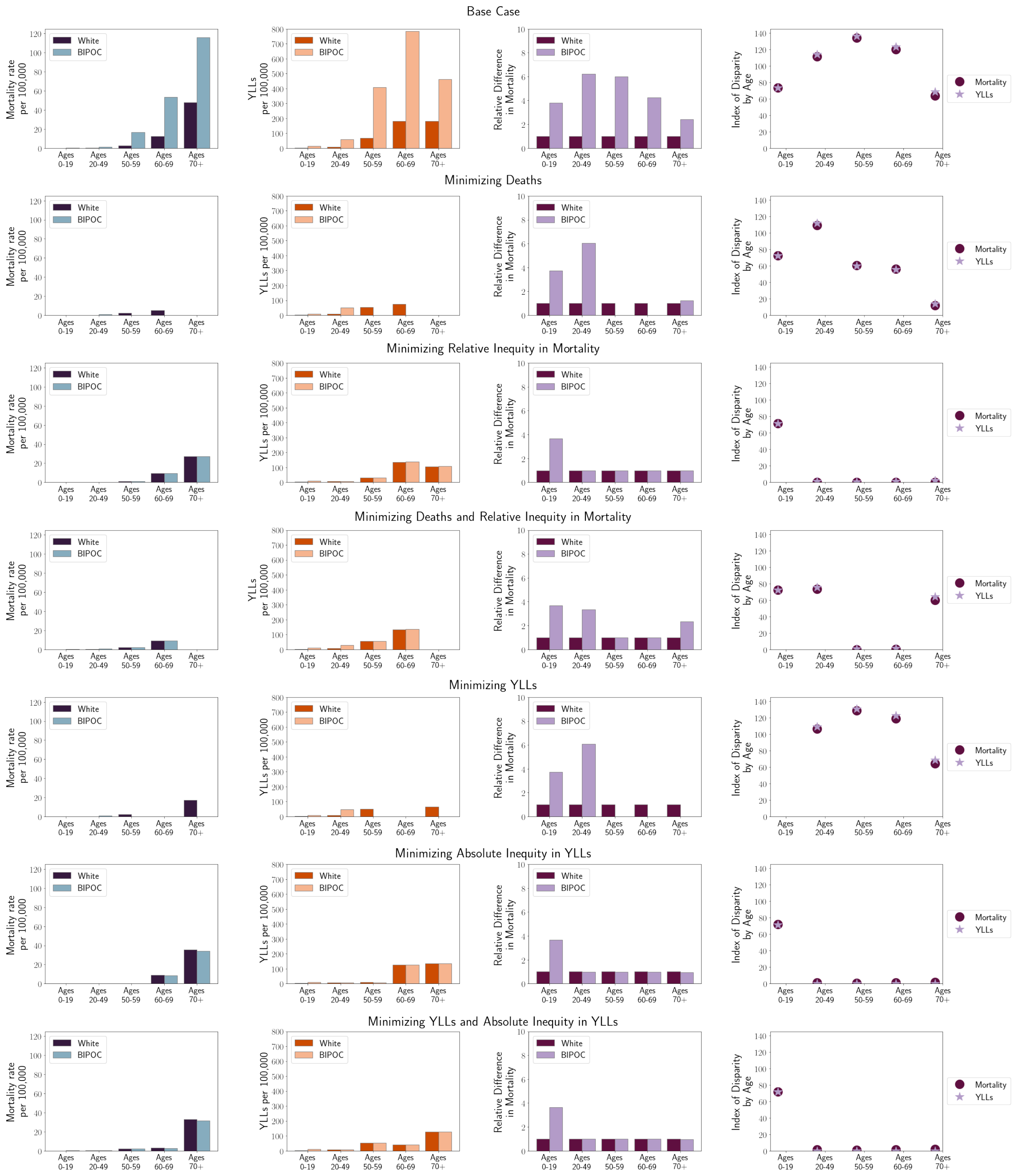


Figure 2: Summary of COVID-19 outcomes by-age when allocating vaccine to 20% of the population. The vaccine allocation strategies that minimize measures of disease burden (rows 2 and 5), inequity measures (rows 3 and 6), and combinations of measures (rows 4 and 7) are evaluated for comparison to the base case (row 1). Outcomes including mortality per 100,000 (column 1), YLLs (column 2), relative disparity in mortality (column 3), and the index of disparity in mortality and YLLs (column 4) are shown.

Table 1: Parameter values used in mathematical model.

| Parameter | Meaning | Value | Source |
|---|---|---|---|
| $1/\sigma$ | mean time from symptom onset to hospitalization | 3.8 days | [2] |
| $1/\gamma_E$ | mean latent period | 3 days | [3], [4] |
| $1/\gamma_P$ | mean pre-symptomatic period | 2 days | [5] |
| $1/\gamma_A$ | mean infectious period of asymptomatic infectives | 6 days | assumed |
| $1/\gamma_I$ | mean infectious period of symptomatic infectives, after developing symptoms | 4 days | [6], [7] |
| $1/\gamma_H$ | mean duration of hospitalization | W:(1/3,1/3,1/4,0.21,1/6),<br>O:(1/3,1/3,1/4,0.22,1/6) | [8] |
| $a$ | proportion of infections that are asymptomatic | age-stratified | fitted |
| $h$ | proportion of symptomatic infections requiring hospitalization | W:(0.002,0.03,0.1,0.17,0.25),<br>O:(0.007,0.12,0.46,0.79,0.68) | [9] |
| $d$ | hospitalization fatality ratio | W:(0.08, 0.03 , 0.11 , 0.17, 0.23),<br>O:(0.08, 0.04, 0.12, 0.13, 0.18) | [10] |
| $r_A$ | relative infectiousness of asymptomatic infections | 0.75 | [8] |
| $r_P$ | relative infectiousness of pre-symptomatic infections | 1 | [11] |
| $r_H$ | relative infectiousness of hospitalized infections | 0 | assumed |
| $R_0$ | basic reproductive number | 3 | [12], [13] |
| $\beta$ | transmission coefficient | calculated | |
| $C$ | contact matrix | - | |
| $N$ | total population | 4060795 | [14] |

Table 2: Deaths per-group (% averted from base case) when minimizing measures of disease burden, measures of inequity, and combinations of measures with enough vaccine to vaccinate 10% of the total population. When minimizing inequity in deaths and when minimizing deaths and inequity, the relative inequity in deaths is used. When minimizing inequity in YLLs and when minimizing YLLs and inequity, the absolute inequity in YLLs is used.

| Age-Group | 0-19 | 20-49 | 50-59 | 60-69 | 70+ |
|---|---|---|---|---|---|
| Base Case: | | | | | |
| White | 0 | 3 | 12 | 58 | 220 |
| Other | 1 | 6 | 14 | 30 | 46 |
| Minimizing Deaths: 61% total deaths averted | | | | | |
| White | 0 (0%) | 3 (0%) | 12 (0%) | 58 (0%) | 56 (75%) |
| Other | 1 (0%) | 6 (0%) | 15 (-7%) | 0 (100%) | 0 (100%) |
| Minimizing Inequity in Deaths: 18% total deaths averted | | | | | |
| White | 0 (0%) | 3 (0%) | 11 (8%) | 57 (2%) | 218 (1%) |
| Other | 0 (100%) | 2 (67%) | 2 (86%) | 7 (77%) | 19(59%) |
| Minimizing Both Deaths and Inequity: 57% total deaths averted | | | | | |
| White | 0 (0%) | 3 (0%) | 11 (8%) | 55 (5%) | 75 (66%) |
| Other | 0 (100%) | 5 (17%) | 2 (86%) | 7 (77%) | 6 (87%) |
| Minimizing YLLs: 42% total deaths averted | | | | | |
| White | 0 (0%) | 2 (33%) | 10 (17%) | 24 (59%) | 183 (17%) |
| Other | 0 (100%) | 5 (17%) | 0 (100%) | 0 (100%) | 0 (100%) |
| Minimizing Inequity in YLLs: 19% total deaths averted | | | | | |
| White | 0 (0%) | 3 (0%) | 11 (8%) | 57 (2%) | 217 (1%) |
| Other | 0 (100%) | 2 (67%) | 2 (86%) | 7 (77%) | 18 (61%) |
| Minimizing Both YLLs and Inequity: 40% total deaths averted | | | | | |
| White | 0 (0%) | 2 (33%) | 10 (17%) | 21 (64%) | 186 (15%) |
| Other | 0 (100%) | 5 (17%) | 2 (86%) | 3 (90%) | 5 (89%) |

Table 3: YLLs per-group (% averted from base case) when minimizing measures of disease burden, measures of inequity, and combinations of measures with enough vaccine to vaccinate 10% of the total population. When minimizing inequity in deaths and when minimizing deaths and inequity, the relative inequity in deaths is used. When minimizing inequity in YLLs and when minimizing YLLs and inequity, the absolute inequity in YLLs is used.

| Age-Group | 0-19 | 20-49 | 50-59 | 60-69 | 70+ |
|---|---|---|---|---|---|
| Base Case: | | | | | |
| White | 3 | 9 | 67 | 180 | 182 |
| Other | 13 | 60 | 408 | 784 | 460 |
| Minimizing Deaths: 62% total YLLs averted | | | | | |
| White | 3 (0%) | 10 (-11%) | 72 (-7%) | 180 (0%) | 47 (74%) |
| Other | 12 (8%) | 65 (-8%) | 429 (-5%) | 0 (100%) | 0 (100%) |
| Minimizing Inequity in Deaths: 59% total YLLs averted | | | | | |
| White | 3 (0%) | 9 (0%) | 65 (3%) | 177 (2%) | 180 (1%) |
| Other | 11 (15%) | 17 (72%) | 66 (84%) | 181 (77%) | 188 (59%) |
| Minimizing Both Deaths and Inequity: 68% total YLLs averted | | | | | |
| White | 3 (0%) | 9 (0%) | 66 (1%) | 170 (6%) | 62 (66%) |
| Other | 11 (15%) | 59 (2%) | 67 (84%) | 174 (78%) | 65 (86%) |
| Minimizing YLLs: 83% total YLLs averted | | | | | |
| White | 3 (0%) | 9 (0%) | 59 (12%) | 74 (59%) | 151 (17%) |
| Other | 11 (15%) | 54 (10%) | 0 (100%) | 0 (100%) | 0 (100%) |
| Minimizing Inequity in YLLs: 59% total YLLs averted | | | | | |
| White | 3 (0%) | 9 (0%) | 65 (3%) | 176 (2%) | 180 (1%) |
| Other | 11 (15%) | 17 (72%) | 65 (84%) | 176 (78%) | 180 (61%) |
| Minimizing Both YLLs and Inequity: 75% total YLLs averted | | | | | |
| White | 3 (0%) | 9 (0%) | 61 (9%) | 66 (63%) | 154 (15%) |
| Other | 11 (15%) | 55 (8%) | 61 (85%) | 66 (92%) | 48 (90%) |

Table 4: Deaths per-group (% averted from base case) when minimizing measures of disease burden, measures of inequity, and combinations of measures with enough vaccine to vaccinate 20% of the total population. When minimizing inequity in deaths and when minimizing deaths and inequity, the relative inequity in deaths is used. When minimizing inequity in YLLs and when minimizing YLLs and inequity, the absolute inequity in YLLs is used.

| Age-Group | 0-19 | 20-49 | 50-59 | 60-69 | 70+ |
|---|---|---|---|---|---|
| Base Case: | | | | | |
| White | 0 | 3 | 12 | 58 | 220 |
| Other | 1 | 6 | 14 | 30 | 46 |
| Minimizing Deaths: 89% total deaths averted | | | | | |
| White | 0 (0%) | 2 (33%) | 9 (25%) | 24 (59%) | 0 (100%) |
| Other | 0 (100%) | 5 (17%) | 0 (100%) | 0 (100%) | 0 (100%) |
| Minimizing Inequity in Deaths: 50% total deaths averted | | | | | |
| White | 0 (0%) | 2 (33%) | 5 (58%) | 44 (24%) | 126 (43%) |
| Other | 0 (100%) | 1 (83%) | 1 (93%) | 5 (83%) | 11 (76%) |
| Minimizing Both Deaths and Inequity: 83% total deaths averted | | | | | |
| White | 0 (0%) | 2 (33%) | 9 (25%) | 43 (26%) | 0 (100%) |
| Other | 0 (100%) | 3 (50%) | 2 (86%) | 5 (83%) | 0 (100%) |
| Minimizing YLLs: 76% total deaths averted | | | | | |
| White | 0 (0%) | 2 (33%) | 9 (25%) | 0 (100%) | 80 (64%) |
| Other | 0 (100%) | 4 (33%) | 0 (100%) | 0 (100%) | 0 (100%) |
| Minimizing Inequity in YLLs: 42% total deaths averted | | | | | |
| White | 0 (0%) | 2 (33%) | 1 (92%) | 41 (29%) | 164 (25%) |
| Other | 0 (100%) | 1 (83%) | 0 (100%) | 5 (83%) | 14 (70%) |
| Minimizing Both YLLs and Inequity: 50% total deaths averted | | | | | |
| White | 0 (0%) | 2 (33%) | 9 (25%) | 13 (78%) | 153 (30%) |
| Other | 0 (100%) | 1 (83%) | 2 (86%) | 2 (93%) | 13 (72%) |

Table 5: YLLs per-group (% averted from base case) when minimizing measures of disease burden, measures of inequity, and combinations of measures with enough vaccine to vaccinate 20% of the total population. When minimizing inequity in deaths and when minimizing deaths and inequity, the relative inequity in deaths is used. When minimizing inequity in YLLs and when minimizing YLLs and inequity, the absolute inequity in YLLs is used.

| Age-Group | 0-19 | 20-49 | 50-59 | 60-69 | 70+ |
|---|---|---|---|---|---|
| Base Case: | | | | | |
| White | 3 | 9 | 67 | 180 | 182 |
| Other | 13 | 60 | 408 | 784 | 460 |
| Minimizing Deaths: 91% total YLLs averted | | | | | |
| White | 3 (0%) | 8 (11%) | 54 (19%) | 75 (58%) | 0 (100%) |
| Other | 10 (23%) | 49 (18%) | 0 (100%) | 0 (100%) | 0 (100%) |
| Minimizing Inequity in Deaths: 73% total YLLs averted | | | | | |
| White | 3 (0%) | 7 (22%) | 32 (52%) | 135 (25%) | 104 (43%) |
| Other | 9 (31%) | 7 (88%) | 32 (92%) | 138 (82%) | 109 (76%) |
| Minimizing Both Deaths and Inequity: 80% total YLLs averted | | | | | |
| White | 3 (0%) | 8 (11%) | 54 (19%) | 133 (26%) | 0 (100%) |
| Other | 10 (23%) | 28 (53%) | 55 (87%) | 137 (83%) | 0 (100%) |
| Minimizing YLLs: 91% total YLLs averted | | | | | |
| White | 3 (0%) | 8 (11%) | 51 (24%) | 0 (100%) | 66 (64%) |
| Other | 10 (23%) | 48 (20%) | 0 (100%) | 0 (100%) | 0 (100%) |
| Minimizing Inequity in YLLs: 74% total YLLs averted | | | | | |
| White | 2 (33%) | 7 (22%) | 8 (88%) | 127 (29%) | 135 (26%) |
| Other | 9 (31%) | 7 (88%) | 8 (98%) | 128 (84%) | 135 (71%) |
| Minimizing Both YLLs and Inequity: 78% total YLLs averted | | | | | |
| White | 3 (0%) | 7 (22%) | 52 (22%) | 41 (77%) | 126 (31%) |
| Other | 10 (23%) | 7 (88%) | 52 (87%) | 41 (95%) | 126 (73%) |
